# COVID-19 (Omicron strain) hospital admissions from a virtual ward – who required further care?

**DOI:** 10.1101/2022.11.04.22281927

**Authors:** Ian Mackay, Megan France, Duncan McAuley, Sean Wing, Mary Wheeldon, Susan Britton, Catherine Todd, Alexandra Pitiris, Leah Barrett-Beck, Elizabeth Rushbrook, Cameron Bennett, Kate McCarthy

**Affiliations:** Royal Brisbane and Women’s Hospital, Herston, Queensland; School of Clinical Medicine, University of Queensland Herston, Queensland; Metro North Hospital and Health Service, Herston, Queensland

**Author notes:** Corresponding author: Ian Mackay, Royal Brisbane and Women’s Hospital, Butterfield St, Herston QLD, Australia 4029. **Declarations:** No funding was received for this study. Declarations of interest: none All relevant ethical guidelines have been followed. Ethics approval was granted by the Royal Brisbane and Women’s Hospital Human Research Ethics Committee (Approval number: EX/2022/QRBW/84877). There was no material reproduced from other sources. This study was not registered on a clinical trials registry.

**Keywords:** COVID-19, Omicron, virtual care, hospital admissions

## Abstract

**Background:** The COVID-19 virtual ward was created to provide care for people at home with COVID-19. Only a small proportion required hospital admission during their care. Given this was a new model of care, little was known about the clinical characteristics and outcomes of patients requiring admission to hospital from the virtual ward platform.

**Aim:** A retrospective observational study with the aim to characterise hospital admission volume, patient epidemiology, clinical characteristics and outcome form a virtual ward in the setting of an Omicron BA.1 and BA.2 outbreak.

**Methods:** A retrospective observational study was performed for all virtual ward patients admitted from 1^st^ January 2022 to 25^th^ March 2022. Patients had to be at least 16 years old to be included. Epidemiological, clinical and laboratory data was reviewed on all patients who required admission to hospital. This was analysed to describe this patient cohort.

**Results:** A total of 7021 patients were cared for on the virtual ward over the study period with 473 referred to hospital for assessment. Twenty-six (0.4%) patients were admitted to hospital during their care on the ward. Twenty-two (84.6%) admissions were COVID-19 related. Fifty three percent of the hospitalised patients were fully vaccinated, and 11 had received prior therapeutics for COVID-19 in the community. There was one ICU admission, and one in-hospital mortality. Shortness of breath was the most common reason for escalation to hospital. Chest pain was the second most common reason and the most common diagnosis after investigation was non-cardiac chest pain that spontaneously resolved.

**Conclusions:** Few patients required admission from the virtual ward in the setting of the Omicron variant (BA.1, BA.2) as a direct result of COVID-19 disease and virtual ward care. Shortness of breath and chest pain were the most common symptoms driving further clinical care.

**What does this paper add to the literature?:** This paper describes the patient cohort with COVID-19 (Omicron variant) who are unable to be cared for by the virtual model of care and required escalation for hospital admission. It assists in health service planning in the setting of large numbers of cases.

## Introduction

The Metro North COVID-19 Virtual Ward was created to provide care for people at home with COVID-19 in South-East Queensland, Central West and Norfolk Island. This covers a population approaching 900,000 and 4157 square kilometres. Within these catchments there 22 public hospitals including 1 quaternary, 1 tertiary and 2 secondary hospitals. The virtual ward structure has been previously described by McCarthy et al (2022) and had constant evolution in response to changing needs of COVID-19 management and demand [1]. Like a traditional hospital ward, the virtual ward had a list of inpatients and was managed by a multidisciplinary team. However, unlike a traditional hospital ward, the virtual ward patients were at home and consultations were provided over the phone or using telehealth.

To provide care the virtual ward needed to be able to recognise and escalate moderate-severe COVID-19 cases, complications of COVID-19 and other medical conditions requiring hospital level care and assessment. In a large multinational observational study, fever, cough, and shortness of breath were the most common symptoms in hospitalised patients for COVID-19 [2]. Atypical symptoms included nausea, vomiting and abdominal pain for people less than 60 and confusion was the most common atypical symptom for those over 60 [2]. A study by O’Malley, Hansjee [3] showed that a virtual ward model was useful in following up high-risk patients discharged from a respiratory ward in a UK hospital. Previous studies have shown that a COVID-19 virtual ward model of care can relieve pressure on hospitals whilst providing safe management of patients at home [3-5].

The aim of this study was to characterise hospital admission volume epidemiology, clinical characteristics and outcome form a virtual ward model of care as a part of service learning in an Omicron BA.1 and BA.2 outbreak. This knowledge will assist future hospital planning and optimise patient care in the virtual ward setting.

## Method

### Study design

A retrospective assessment was performed for all patients admitted to hospital during their virtual ward admission from 1^st^ January 2022 to 25^th^ March 2022. An admission meant that a patient was admitted to a hospital bed under the care of a specialist. Short stay admissions in the emergency department were excluded. The most predominant Omicron strain during this period was BA.1.

Data was obtained from the Virtual Care Stream (Clinical notes system), the Virtual Ward dashboard (Power BI), The Viewer and iEMR. These are online clinical notes systems where a health professional can visualise patient’s previous encounters with public hospitals, including notes and summaries of their previous care. Patients were admitted to the ward as opt-in model based on positive chain reaction (PCR) testing for SARS-CoV-2. Outside the opt in model patients were referred by another practitioner (e.g. the emergency department or the General Practitioner) or self-referred through electronic or phone call platforms. This study was approved by the Royal Brisbane and Women’s Hospital Human Research Ethics Committee (Approval number: EX/2022/QRBW/84877).

### Virtual Ward and Escalation Process

Patients were admitted to the COVID-19 Virtual Ward while in isolation at their home. The Virtual ward included administration, nursing, pharmacy, medical and social work staff. An escalation hotline was available to patients after hours.

In this initial consultation patients were risk stratified based on risk of possible disease progression. All patients received daily phone calls and symptoms were assessed with standardised escalation criteria. Higher risk patients received a pulse oximeter. Additional questions were asked for pregnant patients that screened for any issues with pregnancy. The above system allowed escalation of patients to a Medical Officer for review and then to the emergency department if required. Transport to the nearest emergency department was arranged via an ambulance and the Senior Medical Officer of that department was made aware of the patients expected arrival.

Antiemetics, analgesia, antibiotics and oral antiviral therapies when they became available could be delivered to the patient’s home and were prescribed according to the Australian National Guidelines [6]. Sotrovimab and then EVUSHED (tixagevimab and cilgavimab) at a later point were normally given in the outpatient setting but for some time required hospital presentation until this facility was set up. Key changes in ward strategy are listed in table 1.

**Table 1:**
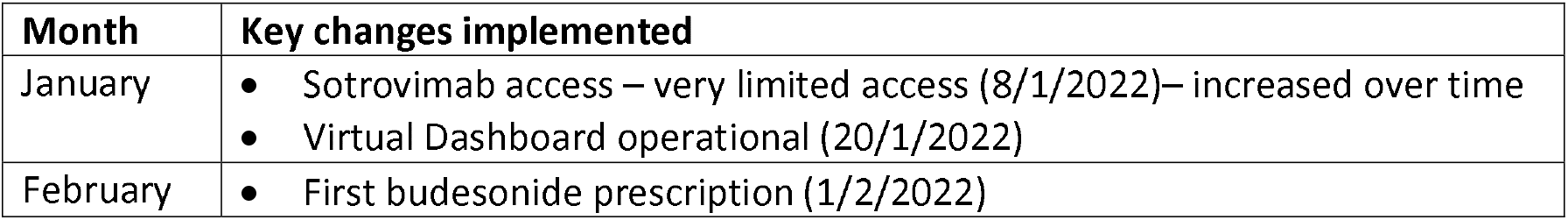
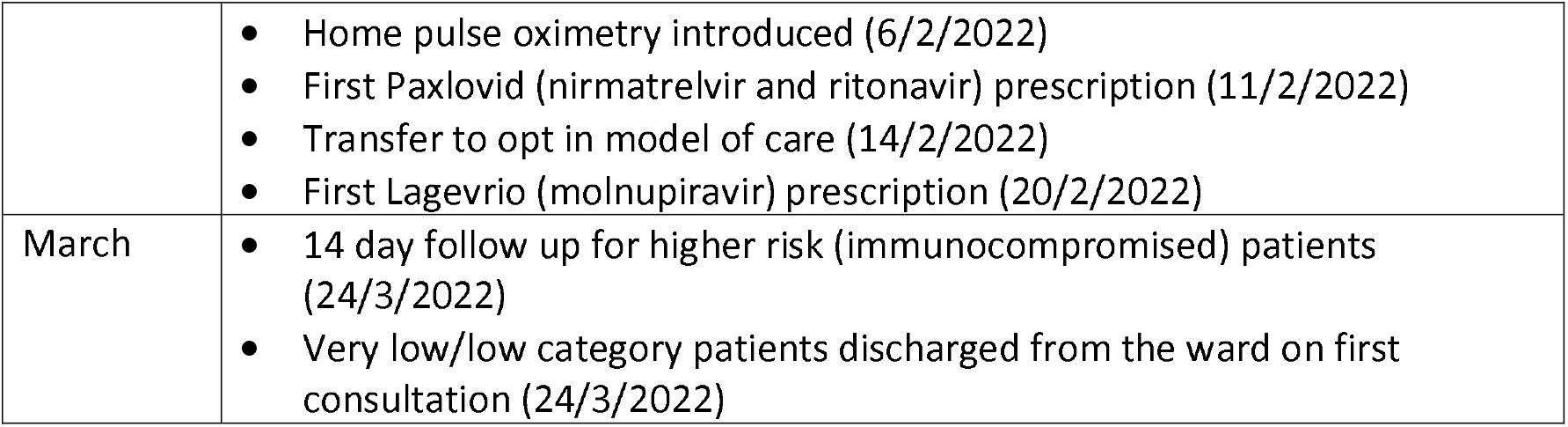
Key changes in the virtual ward

Patients were discharged from the virtual ward after 7 days if they met the national guideline criteria relating to symptom improvement [7].The Virtual Ward implemented a 14 day total follow up, if they had ongoing symptoms, in the heavily immunosuppressed patient cohort such as lung and liver transplant patients.

### Study population

All patients 16 years or over who were admitted to the virtual ward from 1^st^ January 2022 to 25^th^ March 2022 were reviewed. Patients were deemed COVID-19 positive if they had a self-reported or confirmed positive PCR (Polymerase Chain Reaction) test or RAT (Rapid Antigen Test). Patients were excluded from the study as a hospital admission if they were admitted to hospital to receive an intravenous COVID-19 therapeutic only or if they were admitted to a short stay ward associated with any department of that hospital.

### Data Collection

Data was collected on total numbers of patients admitted to the virtual ward, number of consultations, number of patients who attended an emergency department and those admitted to hospital. The patients admitted to hospital were studied in detail including patient demographics, vaccination status, comorbidities (including immunocompromise), COVID-19 testing results, reason for escalation, hospital assessment, pathology results, hospital admission treatments, Virtual Ward disposition and treatment outcome. Vaccination status was defined per the Australian Technical Advisory Group on Immunisation (ATAGI)[8]. Day 0 of COVID-19 was defined as the date the patient was diagnosed or day of first symptoms (not more than 48 hours prior to diagnosis date)[7]. Immunocompromise was defined per ATAGI and included medical conditions such as active haematological malignancy, immunosuppressive therapy, organ transplant with immunosuppressive therapy and primary immunodeficiency syndromes [9]. Mortality data was obtained (in-hospital or within 30 days of discharge) and readmission was defined as a further admission to a hospital (not a short stay unit) within 30 days.

### Data Analysis

The data was analysed using SPSS Statistics 27. Descriptive statistics were expressed as a number (%) and mean or median for continuous variables.

## Results

### Virtual ward admissions, emergency department presentations, and hospital admission rate

A total of 7021 people were actively managed by the virtual ward over the study period. A total of 473 of those patients, 6.7%, attended an emergency department for assessment as a result of escalation of their care by the virtual ward or self-escalation. Of those, 26 patients (0.4% of the total ward patients), were admitted to a hospital for further care (Table 2).

**Table 2:**
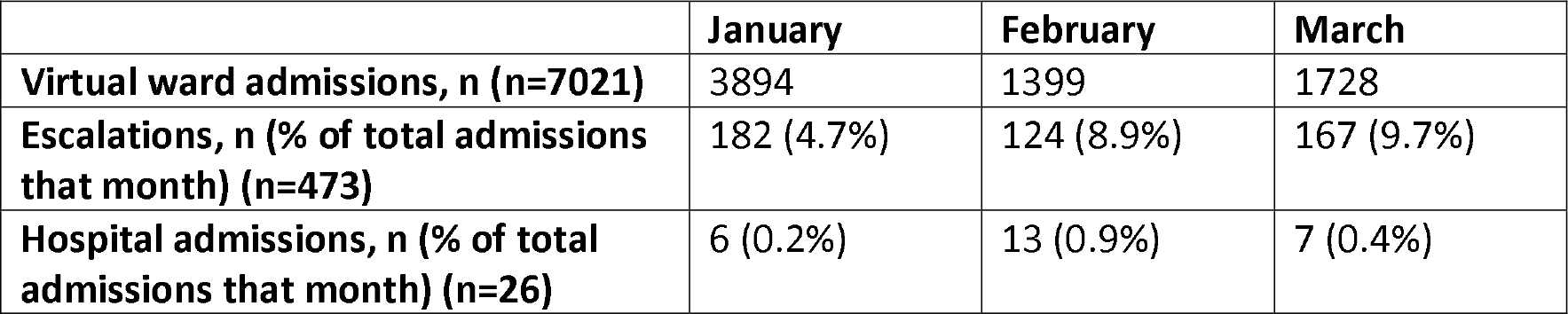
Numbers of patients admitted to the virtual ward, attendance to an emergency department due to escalation, and admitted to a hospital from January to March 2022

A total of 26 patients were admitted to hospital during the period of the study. Patient characteristics are summarised in Table 3. Note this included hospital admissions for all causes, not only COVID-19 admissions.

**Table 3:** Baseline characteristics of Hospitalised patients (n=26)

| Baseline characteristics | All hospitalised patients |
| --- | --- |
| **Median age, years (range)** | 62 (18-89) |
| **Female sex, n (%)** | 17 (65.4) |
| **Vaccination status** |  |
| Fully vaccinated and up to date, n (%) | 11 (42.3) |
| Fully vaccinated, n (%) | 3 (11.5) |
| Partially vaccinated, n (%) | 8 (30.8) |
| Unvaccinated, n (%) | 4 (15.4) |
| **Pregnant, n (%)** | 2 (7.6) |
| **Risk category** |  |
| Very High, n (%) | 13 (50.0) |
| High, n (%) | 11 (42.3) |
| Moderate, n (%) | 2 (7.7) |
| Low/Very Low, n (%) | 0 (0) |
| **Immunocompromise, n (%)** | 5 (19.2) |
| **Median Charlson Comorbidity Index, index (range)** | 2 (0-8) |
| **Other at-risk comorbidities** |  |
| Hypertension, n (%) | 12 (46.2) |
| Diabetes, n (%) | 6 (23.1) |
| Obesity, n (%) | 4 (15.4) |
| Chronic Obstructive Pulmonary Disease, n (%) | 3 (11.5) |
| Asthma (Moderate to Severe), n (%) | 4 (15.4) |
| **Virtual ward treatments** |  |
| Budesonide, n (%) | 7 (26.9) |
| Increased usual inhaled corticosteroid dose, n (%) | 2 (7.7) |
| Sotrovimab, n (%) | 1 (3.8) |
| Molnupiravir, n (%) | 1 (3.8) |
| Paxlovid, n (%) | 0 (0.0) |
| Nil, n (%) | 15 (57.7) |

Table 4 shows the escalation characteristics of the patients who went on to be admitted to hospital. Five (19.2%) patients escalated themselves to hospital for care. Shortness of breath was the most common reason for escalation. The second most common reason was chest pain, the diagnoses of which are listed in Table 5.

**Table IV:** Escalation characteristics (in patients admitted to hospital) (n=26)

|  |  |
| --- | --- |
| **Escalation characteristics** |  |
| **Median COMPASS score (on day of escalation) (n=21)** | 6 |
| **Self-escalated, n (%)** | 5 (19.2) |
| **Median day of COVID-19 illness when escalated, day (range)\*** | 5.5 (2-16) |
| **Escalation symptoms** |  |
| Presence of chest pain, n (%) | 10 (38.5) |
| Presence of shortness of breath, n (%) | 13 (50) |
| Other symptoms | 9 (34.6) |

\*Included day 0

**Table V:**
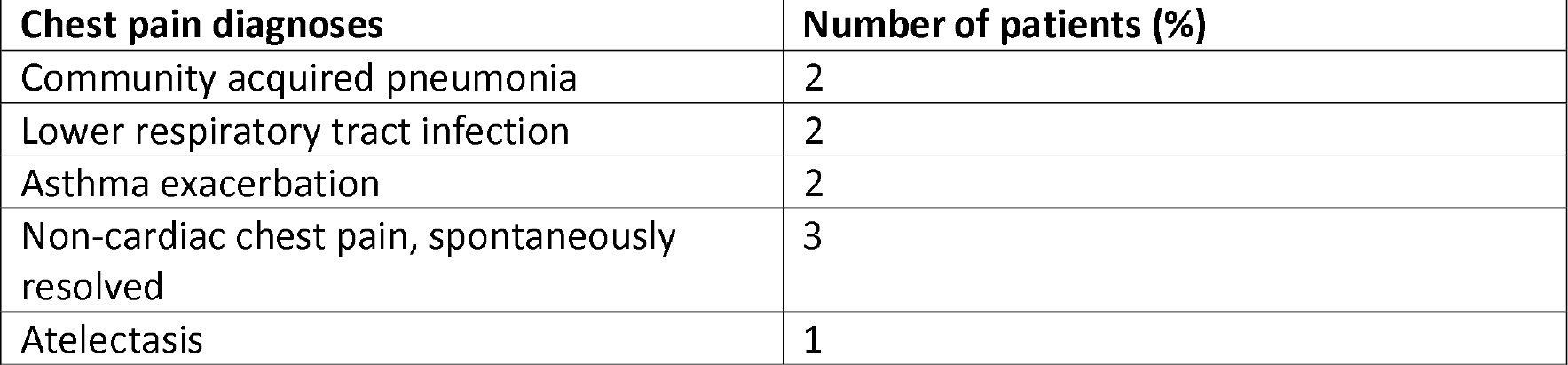
Chest pain diagnoses of patients admitted to hospital (n=10)

Other reasons for escalation included per vaginal bleeding (n=1), flank pain (n=1), dysuria (n=1), desaturation on pulse oximeter (n=2), fevers/severe fatigue (n=1), reduced oral intake (n=2) and tingling of the hands and feet (n=1).

### Admission Characteristics and Outcomes

The summary of the admission data is shown in Table 6.

**Table 6:** Admission summary (n=26)

|  |  |
| --- | --- |
| **Admission characteristics** |  |
| **Median length of Hospital stay, days (range)** | 3 (0-21) |
| **ICU admissions, n (%)** | 1 (3.8) |
| **COVID-19 Illness Severity** |  |
| Mild, n (%) | 6 (23.1) |
| Moderate, n (%) | 10 (38.5) |
| Severe, n (%) | 7 (26.9) |
| Critical, n (%) | 2 (7.7) |
| **COVID-19 related admissions, n (%)** | 22 (84.6) |
| **COVID-19 Treatments during hospitalisation** |  |
| Budesonide/Other Inhaled Corticosteroid, n (%) | 5 (19.2) |
| Dexamethasone, n (%) | 11 (42.3) |
| Oral steroids, n (%) | 10 (38.5) |
| Remdesivir, n (%) | 1 (3.8) |
| Baricitinib, n (%) | 2 (7.7) |
| Oxygen therapy, n (%) | 10 (38.5) |
| **Admission Outcome** |  |
| In-hospital mortality, n (%) | 1 (3.8) |
| Discharge to home, n (%) | 25 (96.2) |
| **30-day outcome** |  |
| Readmission within 30 days, n (%) | 1 (4) |
| 30-day mortality, n (%) | 0 (0) |

There were no instances of cardiac chest pain or pulmonary embolus substantiated by relevant imaging/pathology results. One patient was admitted for an asthma exacerbation and tested negative for COVID-19, having already completed the 7-day isolation period and therefore was classed as an unrelated admission. Admissions not directly related to COVID-19 included antepartum haemorrhage (n=1), Epstein Barr Virus infection (n=1) and urinary tract infection (n=1).

## Discussion

A total of 7021 patients were cared for on the virtual ward over the study period. Only 0.4% of those patients were admitted to hospital. There were 473 (6.7%) patients escalated to the emergency department for further assessment. There was no reduction in the number of patients admitted to hospital per month for the three months that this data was collected indicating that the changes in process of the virtual ward did not change admission/escalation rates.

One reason for the low admission rate is potentially due to the mass vaccination rollout as this has been shown to significantly reduce risk of hospital admission [10]. Previous studies have shown that there is an increase in hospital mortality with floods of patients indicating the need for streamlined pathways for admission [11]. Studies have also shown that the COVID-19 pandemic has decreased hospital presentations, for example in three hospitals in America showing reduced psychiatric presentations and hospitalisations [12]. There was also a decreased in presentations of Chronic Obstructive Pulmonary Disease exacerbations due to physical and behavioural measures taken to limit COVID-19 transmission [13].

Fifty three percent of the patients admitted to hospital from the virtual ward were fully vaccinated. This is significantly lower than the population vaccination rate at this time which was gradually increasing and by 20 March 2022 was greater than 80% of the community population [14]. Almost 60% of the cohort were not able to receive therapeutics due to their unavailability or less commonly patent refusal. Thus, therapeutics did not pay a major role in prevention of hospital admission.

Shortness of breath was the most common reason for a Medical Officer to escalate a patient for in-hospital assessment, despite the availability of pulse oximeters. As this was a new unvalidated process at this time clinician discretion was utilised as to whether the readings changed patient care and if a patient was kept at home when experiencing this symptom with normal oximetry.

Chest pain was a common complaint amongst patients admitted to hospital, however there were no sinister causes of the chest pain found on further investigation. The workup for these patients included basic bloods (full blood count, electrolytes, liver function, kidney function), troponin, chest x-ray and an electrocardiogram. Three of the 10 patients with chest pain had a d-dimer and one had a CT pulmonary angiogram. The most common diagnosis amongst this cohort was non-cardiac chest pain that spontaneously resolved but also included lower respiratory tract infection and asthma exacerbation. Knowing this would allow the virtual ward to potentially reduce the number of escalations to hospital in patients with chest pain in the future. This represents greater familiarity of disease manifestations form the Omicron variant.

In-hospital mortality for this group was 3.8% (n=1) and ICU admissions was 3.8% (n=1). This is largely different to the epidemiology and clinical characteristics of COVID-19 in Wuhan in 2019 with the alpha variant where there were 138 hospitalised patients with COVID-19 pneumonia, with 26% needing ICU treatment and a mortality of 4.3% [15]. Previous studies have shown that the delta outbreak in unvaccinated population would lead to a greater burden on the health care system [16]. This study has shown that the burden on the hospital system was very low with the implementation of the virtual ward.

Study limitations included that, in the latter half of the study, the model of care was an “opt in” one and thus patients admitted to the virtual ward were self-selected. The change in patients’ management that occurs with greater familiarity with disease manifestations of a new variant and greater familiarity with technology as it is introduced such as pulse oximetry. Also, availability of therapeutics increased in the later half of the study period.

## Conclusions

From the virtual ward setting hospital presentations to the emergency department and for admission was a small percentage of the cohort. This was in the setting of a vaccination rate of 57%, limited therapeutics for most of the study period and when Omicron (BA.1/BA.2) was the predominant strain. Shortness of breath and chest pain were the most common symptoms resulting in hospital admission.

## Supporting information

Research Reporting Checklist

## Data Availability

All data produced in the present work are contained in the manuscript.

## Acknowledgements

Nil

## Notes

### Competing Interest Statement

The authors have declared no competing interest.

### Funding Statement

This study did not receive any funding.

### Author Declarations

Royal Brisbane and Womens Hospital Human Research Ethics Committee of Royal Brisbane and Womens Hospital gave ethical approval for this work.

